# Who Gets Help? Analyzing Disparities in Diagnosis, Treatment, and Care Seeking of Anxiety and Depression Among Women in Bangladesh

**DOI:** 10.1101/2025.05.07.25327186

**Authors:** Isna Haque Sheoti, Md. Zakiul Alam

**Affiliations:** Department of Population Sciences, University of Dhaka, Dhaka-1000, Bangladesh; Department of Population, Family and Reproductive Health, Johns Hopkins Bloomberg School of Public Health, Johns Hopkins University, Maryland-21205, USA

**Keywords:** Diagnosis, Treatment, Care-seeking, Anxiety, Depression, Anxiety in Bangladesh, Depression in Bangladesh

## Abstract

**Introduction:** Despite the availability of various treatments for anxiety and depression, the diagnosis, treatment, and care-seeking are very low in Bangladesh, which can have effects on both individual and societal levels. We explored the situation and factors associated with the diagnosis, treatment, and care-seeking of anxiety and depression among reproductive-aged women in Bangladesh.

**Methodology:** We utilized data from the Bangladesh Demographic and Health Survey (BDHS), 2022. Diagnosis (ever told by a healthcare provider), treatment (took prescribed medicine), and care-seeking (who have ever sought help) were outcome variables. We used log-binomial logistic regression to explore the factors associated with diagnosis, treatment, and care-seeking of anxiety and depression.

**Findings:** Among all respondents, 3.39% were diagnosed with anxiety, 0.56% were diagnosed with depression by a provider, and only 2.0% received treatment, with 7.9% seeking help from a doctor or medical personnel. Compared to women aged 15-19, diagnosis and care-seeking were significantly higher among women aged 20-49. Significant regional variations were observed for treatment and care seeking. Women with a positive attitude towards wife beating had significantly higher levels of diagnosis (PR=1.28, CI: 1.06, 1.45, p=0.008), treatment (PR=1.50, CI: 1.17, 1.92, p=0.001), and care-seeking (PR=1.52, CI: 1.34, 1.72, p<0.001) than others. Middle class (PR=1.19, CI: 1.00, 1.41, p=0.046) and working women (PR=1.22, CI: 1.09, 1.38, p=0.001) had a higher, and women who didn’t use the internet (PR=0.67, CI: 0.59, 0.77, p<0.001) had a lower prevalence of care-seeking.

**Conclusion:** Policy actions should be taken to reduce the stigma towards asking for and receiving help for common mental health disorders and create a specialized department in government hospitals. Internet and social media can be used strategically for this, and special focus should be given to women suffering from intimate partner violence.

## Introduction

Anxiety and depression are the most common mental health disorders in the world.[1,2] Around 4% of the world’s population suffers from anxiety, and 5% suffers from depression.[1,2] Despite its higher prevalence, the treatment and care-seeking of anxiety and depression were low globally, especially in low-income countries.[3] There also exists a gap between diagnosis, care seeking, and receiving adequate treatment.[3,4] Even evidence from higher-income countries suggested that only 25% of people suffering from anxiety problems receive proper treatment.[3–5] A higher number of women face anxiety and depression than men.[1,2] In Bangladesh, around 20% of reproductive-aged women face anxiety, and 5% of women face depression.[6]

There are many treatment options available for anxiety disorders. For anxiety, selective serotonin reuptake and cognitive-behavioral therapy were found to be effectual.[7] Various scientific studies often suggest a combination of psychological therapy and pharmacotherapy.[8] Similarly, there are several treatment options available for depression which can assist to lessen the symptoms and severity of depression.[9] One option is psychotherapy, which is also called therapy or counseling, prioritizing thoughts, issues in life, and feelings of a patient.[9] Alternatively, there is a scope of treatment with medications commonly known as antidepressants.[9] Healthcare professionals often suggest lifestyle changes simultaneously with treatment for treating both anxiety and depression. [8–10]

If anxiety and depression are not diagnosed and appropriately treated, it can have effects on both individual and societal levels. [11,12] At the individual level, untreated anxiety and depression can have consequences like reduced productivity, disability, heightened risk of self-harm and suicide, and overall a decreased quality of life. [11–13] Individual’s loss of productive time eventually affects the total economic situation of a country. [11] The economic loss of anxiety was the same for dementia in European countries, which portrays the severity of it. [11] The untreated anxiety and depression can have distinct effects on reproductive-aged women as studies presented a surging risk of preterm birth, spontaneous abortion, perinatal depression, and low birth weight. [14,15]

The above discussion shows the necessity to explore the situation of diagnosis, care seeking, and treatment of anxiety and depression among reproductive-aged women. Very few studies were conducted in Bangladesh on this aspect.[16–20] Those studies mainly focused on the pattern of admission and discharge in a hospital, motivation and barriers in care seeking, care seeking behavior, and treatment gap among the adult population, cancer patients, COVID-19 situation, and rural population.[16–20] Only one content analysis was conducted concerning women’s mental health care-seeking behavior. [21] Under these circumstances, our study aims to explore the situation of diagnosis, treatment, and care-seeking behavior of anxiety and depression and factors associated with them among reproductive-aged women (aged 15-49) in Bangladesh by using recent Bangladesh Demographic and Health Survey (BDHS-2022) data.

### Data and methods Data source

This study used data from BDHS-2022, a nationally representative cross-sectional survey.[6] The detailed methodology is available elsewhere.[6] The study used two-stage stratified cluster sampling to collect data on reproductive women’s mental health and well-being. Firstly, they expected to collect data from 30,375 households in 675 enumeration areas (EAs). Later, one enumeration area in Cox’s Bazar was excluded for safety reasons. For that, data was collected only from 674 EAs. Among 45 households in each EA, 30 were selected for a long questionnaire, which included the mental health and well-being section. All 20,029 women were asked questions about diagnosis and treatment. Among them, those who reported any symptoms of anxiety or depression in the preceding two weeks were asked questions about care-seeking with a sample size of 17,192.

### Variables of study Dependent variables

This study has three main dependent variables: diagnosis, treatment, and care-seeking of anxiety and depression. BDHS measured these variables in the long questionnaire. They operationalized diagnosis as ‘percentage of ever-married women aged 15–49 who have ever been told by a health care provider that they have anxiety or depression’. Treatment was defined as ‘percentage of ever-married women aged 15-49 who took medicine prescribed by a health care provider for anxiety or depression in the 2 weeks preceding the survey’. Finally, care-seeking was defined as ‘Among women with any symptoms of anxiety or depression in the 2 weeks preceding the survey, the percentage who have ever sought help’.

### Independent variables

The independent variables employed in this study are age, place of residence, division, wealth index, highest educational level, working status, participation in decision-making, having an account in a bank or other financial institution, religion, attitudes toward wife beating, husband’s level of education, exposed to any mass media, and ever used internet (presented in Table 1). The wealth index measured socioeconomic status and divided into five economic quintiles (poorest, poorer, middle, richer, and richest) based on principal component analysis. Exposure to mass media was measured by whether women had access to any TV, radio, or newspaper. By using four types of information provided by DHS, we measured women’s participation in household decision-making: 1) the person who normally decides on large household decisions, 2) the person who generally makes decisions on family or relatives’ visits, 3) the person who determines typically expenditures of household earnings, and 4) the person who generally makes decisions on the respondent’s health care. Finally, if all the decisions were made by the respondent alone or together with the husband, it was defined as ‘yes,’ and if all the decisions were made by the husband alone or others, it was defined as ‘no.’ Again, attitude towards wife beating was measured as ‘yes’ if respondents responded beating is justified in any of the following items: 1) wife neglects the children, 2) wife goes out without telling husband, 3) wife refuses to have sex with husband, and 4) wife argues with husband. It was measured as ‘no’ if they responded negatively to all items.

**Table 1:** Sample characteristics by diagnosis, treatment, and care-seeking of anxiety/depression.

|  | Diagnosis |  | Treatment |  | Care seeking |  |
| --- | --- | --- | --- | --- | --- | --- |
|  | No | Yes | No | Yes | No | Yes |
| Variables | 18,511 (92.4%) | 1,518 (7.6%) | 19,621 (98.0%) | 407 (2.0%) | 15,149 (88.1%) | 2,043 (11.9%) |
| **Age in 5-year groups** |  |  |  |  |  |  |
| 15-19 | 1,653 (8.9%) | 76 (5.0%) | 1,704 (8.7%) | 25 (6.1%) | 1,215 (8.0%) | 121 (5.9%) |
| 20-24 | 3,131 (16.9%) | 159 (10.5%) | 3,257 (16.6%) | 33 (8.0%) | 2,425 (16.0%) | 273 (13.4%) |
| 25-29 | 3,262 (17.6%) | 261 (17.2%) | 3,459 (17.6%) | 65 (15.8%) | 2,637 (17.4%) | 346 (16.9%) |
| 30-34 | 3,198 (17.3%) | 239 (15.7%) | 3,371 (17.2%) | 66 (16.1%) | 2,651 (17.5%) | 367 (18.0%) |
| 35-39 | 3,046 (16.5%) | 299 (19.7%) | 3,266 (16.6%) | 79 (19.3%) | 2,557 (16.9%) | 388 (19.0%) |
| 40-44 | 2,302 (12.4%) | 244 (16.1%) | 2,469 (12.6%) | 77 (18.9%) | 1,979 (13.1%) | 290 (14.2%) |
| 45-49 | 1,918 (10.4%) | 241 (15.9%) | 2,095 (10.7%) | 64 (15.7%) | 1,684 (11.1%) | 258 (12.6%) |
| **Respondent's current age** | 31.641 (8.914) | 34.354 (8.747) | 31.787 (8.926) | 34.699 (8.635) | 32.083 (8.897) | 33.189 (8.687) |
| **Division** |  |  |  |  |  |  |
| Barishal | 1,122 (6.1%) | 77 (5.1%) | 1,173 (6.0%) | 26 (6.4%) | 1,044 (6.9%) | 66 (3.2%) |
| Chattogram | 3,414 (18.4%) | 335 (22.1%) | 3,643 (18.6%) | 106 (26.0%) | 2,756 (18.2%) | 445 (21.8%) |
| Dhaka | 4,799 (25.9%) | 281 (18.5%) | 4,996 (25.5%) | 84 (20.7%) | 3,869 (25.5%) | 460 (22.5%) |
| Khulna | 2,155 (11.6%) | 233 (15.4%) | 2,357 (12.0%) | 31 (7.7%) | 1,742 (11.5%) | 315 (15.4%) |
| Mymensingh | 1,459 (7.9%) | 68 (4.5%) | 1,496 (7.6%) | 31 (7.6%) | 1,097 (7.2%) | 117 (5.7%) |
| Rajshahi | 2,330 (12.6%) | 295 (19.4%) | 2,557 (13.0%) | 68 (16.8%) | 1,924 (12.7%) | 304 (14.9%) |
| Rangpur | 2,131 (11.5%) | 159 (10.5%) | 2,255 (11.5%) | 36 (8.8%) | 1,768 (11.7%) | 241 (11.8%) |
| Sylhet | 1,100 (5.9%) | 69 (4.5%) | 1,144 (5.8%) | 25 (6.0%) | 950 (6.3%) | 95 (4.6%) |
| **Type of place of residence** |  |  |  |  |  |  |
| Urban | 5,293 (28.6%) | 407 (26.8%) | 5,600 (28.5%) | 100 (24.6%) | 4,307 (28.4%) | 540 (26.5%) |
| Rural | 13,217 (71.4%) | 1,111 (73.2%) | 14,021 (71.5%) | 307 (75.4%) | 10,842 (71.6%) | 1,502 (73.5%) |
| **Highest educational level** |  |  |  |  |  |  |
| No education | 2,503 (13.5%) | 250 (16.4%) | 2,692 (13.7%) | 61 (14.9%) | 2,237 (14.8%) | 241 (11.8%) |
| Primary | 4,796 (25.9%) | 418 (27.5%) | 5,097 (26.0%) | 117 (28.7%) | 4,061 (26.8%) | 527 (25.8%) |
| Secondary | 8,676 (46.9%) | 682 (45.0%) | 9,177 (46.8%) | 182 (44.6%) | 6,904 (45.6%) | 996 (48.8%) |
| Higher | 2,535 (13.7%) | 168 (11.1%) | 2,655 (13.5%) | 48 (11.8%) | 1,948 (12.9%) | 279 (13.6%) |
| **Education in single years** | 6.758 (4.193) | 6.218 (4.225) | 6.723 (4.195) | 6.394 (4.320) | 6.554 (4.203) | 6.977 (4.172) |
| **Wealth of the household** |  |  |  |  |  |  |
| Poorest | 3,336 (18.0%) | 247 (16.3%) | 3,518 (17.9%) | 65 (15.9%) | 2,810 (18.6%) | 342 (16.8%) |
| Poorer | 3,686 (19.9%) | 341 (22.5%) | 3,944 (20.1%) | 83 (20.4%) | 3,095 (20.4%) | 399 (19.6%) |
| Middle | 3,799 (20.5%) | 337 (22.2%) | 4,062 (20.7%) | 74 (18.1%) | 3,097 (20.4%) | 451 (22.1%) |
| Richer | 3,891 (21.0%) | 298 (19.6%) | 4,098 (20.9%) | 91 (22.3%) | 3,141 (20.7%) | 430 (21.0%) |
| Richest | 3,799 (20.5%) | 295 (19.4%) | 3,999 (20.4%) | 95 (23.3%) | 3,006 (19.8%) | 420 (20.6%) |
| **Respondent currently working** |  |  |  |  |  |  |
| No | 12,619 (68.2%) | 998 (65.7%) | 13,334 (68.0%) | 283 (69.4%) | 10,327 (68.2%) | 1,266 (62.0%) |
| Yes | 5,891 (31.8%) | 520 (34.3%) | 6,287 (32.0%) | 125 (30.6%) | 4,822 (31.8%) | 776 (38.0%) |
| **Has an account in a bank or other financial institution** |  |  |  |  |  |  |
| No | 14,865 (80.3%) | 1,164 (76.7%) | 15,734 (80.2%) | 295 (72.5%) | 12,173 (80.4%) | 1,540 (75.4%) |
| Yes | 3,645 (19.7%) | 354 (23.3%) | 3,887 (19.8%) | 112 (27.5%) | 2,976 (19.6%) | 503 (24.6%) |
| **Participation in decision-making** |  |  |  |  |  |  |
| No | 5,691 (30.7%) | 428 (28.2%) | 5,983 (30.5%) | 136 (33.4%) | 4,615 (30.5%) | 615 (30.1%) |
| Yes | 12,820 (69.3%) | 1,090 (71.8%) | 13,638 (69.5%) | 271 (66.6%) | 10,534 (69.5%) | 1,428 (69.9%) |
| **Attitudes toward wife beating** |  |  |  |  |  |  |
| Yes | 2,820 (15.2%) | 295 (19.4%) | 3,027 (15.4%) | 88 (21.7%) | 2,321 (15.3%) | 460 (22.5%) |
| No | 15,691 (84.8%) | 1,223 (80.6%) | 16,594 (84.6%) | 319 (78.3%) | 12,828 (84.7%) | 1,583 (77.5%) |
| **Husband's level of education** |  |  |  |  |  |  |
| No education | 3,885 (21.0%) | 401 (26.4%) | 4,189 (21.3%) | 98 (23.9%) | 3,391 (22.4%) | 420 (20.6%) |
| Primary | 5,268 (28.5%) | 400 (26.4%) | 5,571 (28.4%) | 98 (24.1%) | 4,346 (28.7%) | 565 (27.7%) |
| Secondary | 6,067 (32.8%) | 472 (31.1%) | 6,397 (32.6%) | 142 (34.8%) | 4,876 (32.2%) | 663 (32.5%) |
| Higher | 3,290 (17.8%) | 245 (16.1%) | 3,465 (17.7%) | 70 (17.2%) | 2,535 (16.7%) | 394 (19.3%) |
| **Religion** |  |  |  |  |  |  |
| Others | 1,807 (9.8%) | 114 (7.5%) | 1,877 (9.6%) | 45 (10.9%) | 1,431 (9.4%) | 188 (9.2%) |
| Muslim | 16,703 (90.2%) | 1,404 (92.5%) | 17,744 (90.4%) | 363 (89.1%) | 13,718 (90.6%) | 1,854 (90.8%) |
| **Exposed to any mass media** |  |  |  |  |  |  |
| No | 7,781 (42.0%) | 651 (42.9%) | 8,268 (42.1%) | 164 (40.2%) | 6,452 (42.6%) | 795 (38.9%) |
| Yes | 10,729 (58.0%) | 867 (57.1%) | 11,353 (57.9%) | 244 (59.8%) | 8,697 (57.4%) | 1,248 (61.1%) |
| **Ever used internet** |  |  |  |  |  |  |
| No | 12,904 (69.7%) | 1,096 (72.2%) | 13,717 (69.9%) | 283 (69.4%) | 10,841 (71.6%) | 1,296 (63.5%) |
| Yes | 5,607 (30.3%) | 422 (27.8%) | 5,904 (30.1%) | 125 (30.6%) | 4,308 (28.4%) | 746 (36.5%) |
| **Total Sample** | 20,029 |  | 20,029 |  | 17,192 |  |

### Statistical analysis

First, we analyzed sample characteristics by outcomes (diagnosis, treatment, and care-seeking) **(Table 1)**. Rao and Scott’s corrected Chi-square test was used to measure the association.[22] To explore the factors associated with diagnosis, treatment, and care-seeking of anxiety and depression, we used log-binomial regression. Logistic regression is better suited and extensively used in literature for binary outcomes from cross-sectional data. However, in case of more than 5% prevalence, the odds ratio (OR) overestimates the prevalence ratio (PR). Moreover, log-binomial regression is more suitable for communication than logistic regression.[23] Thus, we used log-binomial regression and ran two models (adjusted and unadjusted). We checked multicollinearity with the variance inflation factor (VIF) using linear regression. The logistic regression model was used to test goodness-of-fit (GOF). We found our model to fit the data. We presented PR, 95% confidence interval (CI), and p-value. All the analyses of this study were conducted using Stata 18, considering survey weights and a complex design.

### Ethics statement

Our study used secondary data from the Bangladesh Demographic and Health Survey 2022. For this reason, we didn’t need any separate IRB approval. DHS survey is conducted by the Ministry of Health and Family Welfare and The National Institute of Population Research and Training (NIPORT). It was implemented by the Mitra and Associates of Bangladesh. ICF International of Calverton, Maryland, USA, gave technical assistance. The survey collected verbal informed consent by using an informed consent statement and NIPORT took ethical approval from the Bangladesh Medical Research Council (BMRC) for the survey. The data set is available at https://dhsprogram.com/data. The total ethics statement can be found in the report. [6]

## Results

Figure 2. presents the situation of diagnosis and treatment of anxiety and depression among respondents. Only 2.03% of respondents took medication or treatment provided by a doctor or health care worker. In the case of diagnosis, 3.39% of respondents were diagnosed with anxiety, 0.56% were diagnosed with depression, and 3.62% were diagnosed with both by a provider.

**Figure 1:**
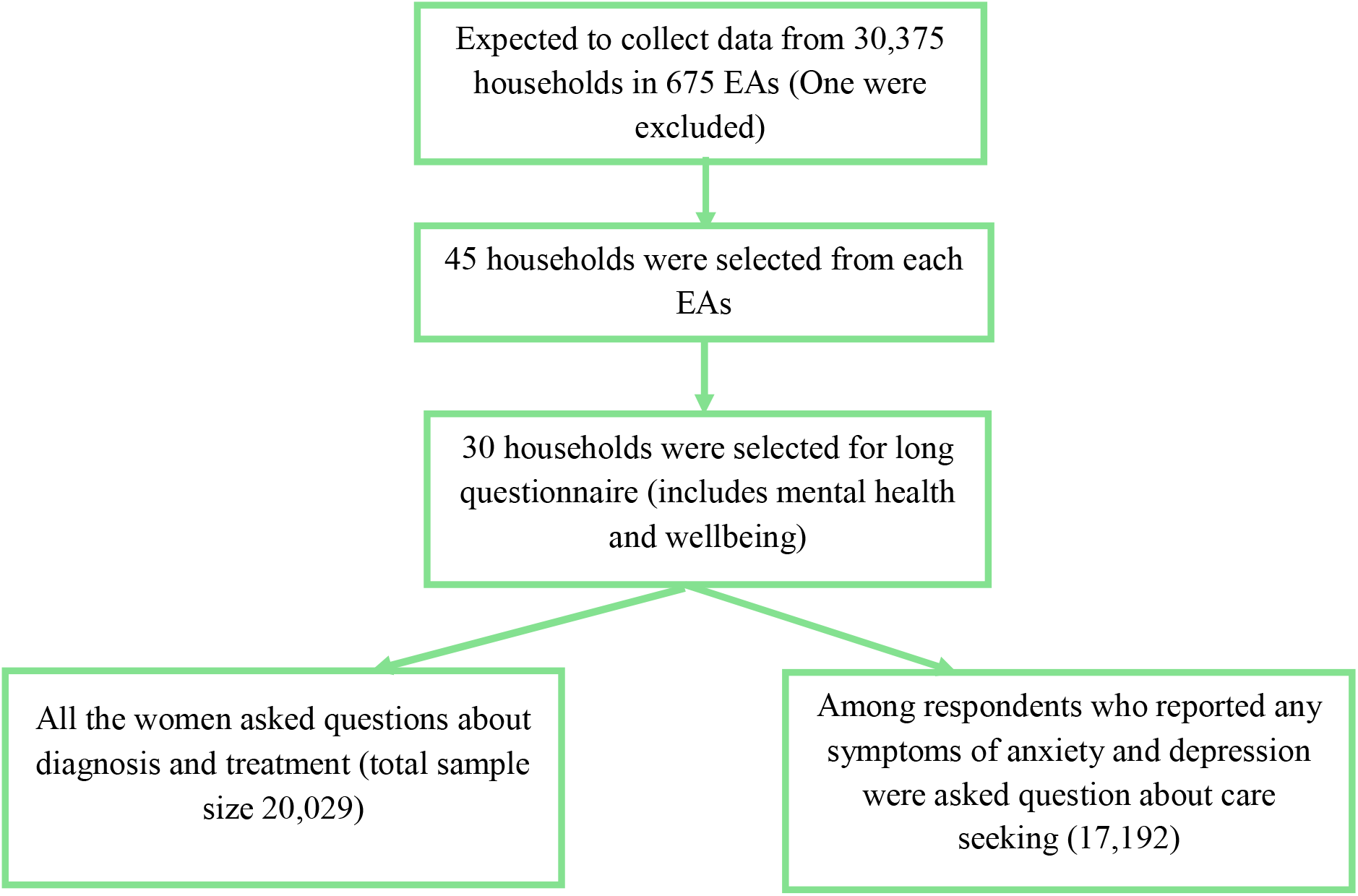
Sample selection process for measurement of diagnosis, treatment, and care-seeking of anxiety and depression

**Figure 2:**
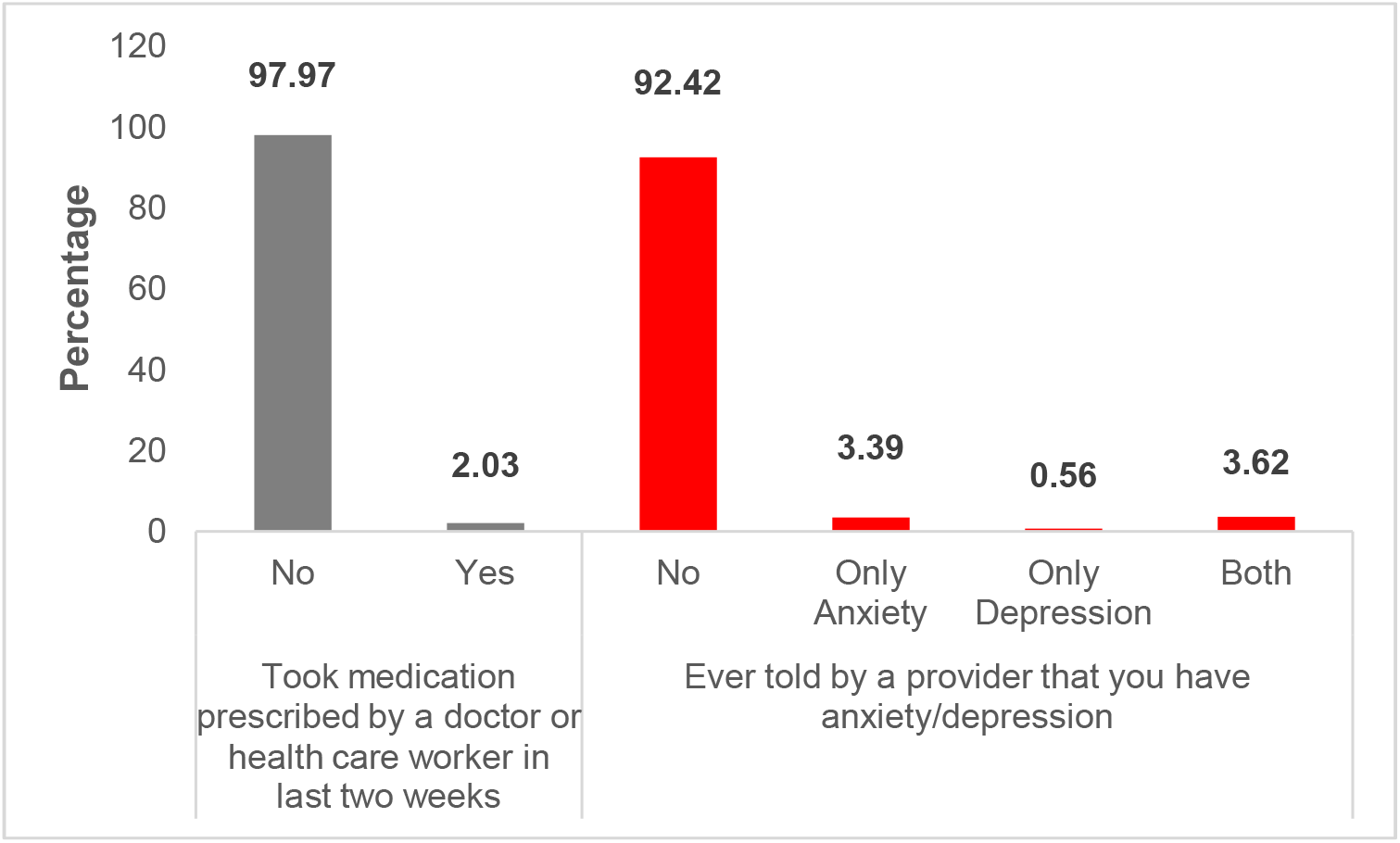
Situation of Diagnosis and treatment of anxiety and depression

Figure 3. presents the source of care-seeking among respondents. Most respondents sought care from their family members (36.08%), followed by current or former spouse or partner (32.02%).

**Figure 3:**
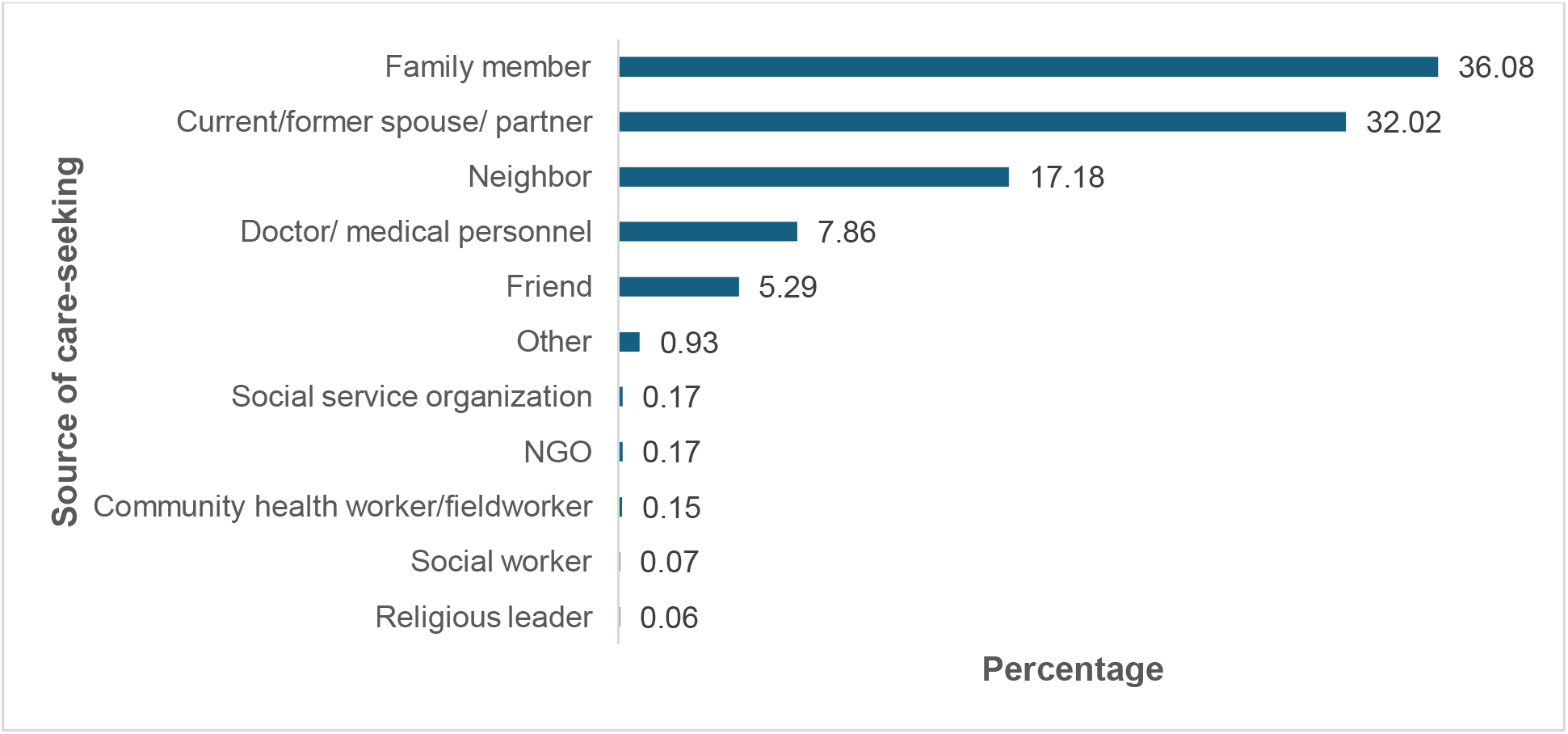
Source of care-seeking among respondents

While 17.18% of respondents sought help from neighbors, only 7.86% sought help from doctors/medical personnel.

Table 1. presents sample characteristics by diagnosis, treatment, and care-seeking of anxiety and depression among respondents. Respondents who received the diagnosis (19.7%), treatment (19.3%), and care-seeking (19.0%) were mostly from the 35-39 years of age. Respondents receiving diagnosis (45.0%), treatment (44.6%), and care (48.8%) were highest from the secondary educational level. While respondents who received diagnosis, treatment, and care were highest from women not working and women who didn’t justify wife beating. On the contrary, respondents who received the diagnosis (92.5%), treatment (89.1%), and care-seeking (90.8%) were predominantly Muslim.

**Table 2** presents factors associated with diagnosis of anxiety and depression among participants using log-binomial regression. Women aged 25 or higher had significant differences with women aged 15-19 in their prevalence of diagnosis. Compared to women aged 15-19, the prevalence of diagnosis was significantly higher among those aged 25-29 (PR:1.74, CI: 1.29, 2.35, p<0.001), 30-34 (PR:1.60, CI: 1.18, 2.18, p=0.003), 35-39 (PR:2.03, CI: 1.52, 2.71, p<0.001), 40-44 (PR:2.18, CI: 1.59, 2.99, p<0.001), and 45-49 (PR=2.52, CI: 1.86, 3.42, p<0.001). Women living in Khulna (PR=2.17, CI: 1.49, 3.16, p<0.001) and Rajshahi (PR=2.44, CI: 1.66, 3.58, p<0.001) had a significantly higher prevalence of diagnosis of anxiety and depression than other divisions. Lastly, women with a positive attitude towards wife beating had significantly higher levels of diagnosis (PR=1.28, CI: 1.06, 1.45, p=0.008) than women with a negative attitude.

**Table 2:** Factors associated with diagnosis of anxiety and depression using log-binomial regression.

| Variables | Unadjusted model |  | Adjusted model |  |
| --- | --- | --- | --- | --- |
|  | Prevalence ratio [95% CI] | p-value | Prevalence ratio [95% CI] | p-value |
| **Age of the respondent** |  |  |  |  |
| 20-24 | 1.09[0.82,1.46] | 0.524 | 1.13[0.84,1.51] | 0.410 |
| 25-29 | 1.69[1.26,2.26] | <0.001 | 1.74[1.29,2.35] | <0.001 |
| 30-34 | 1.58[1.18,2.13] | 0.002 | 1.60[1.18,2.18] | 0.003 |
| 35-39 | 2.03[1.54,2.68] | <0.001 | 2.03[1.52,2.71] | <0.001 |
| 40-44 | 2.18[1.62,2.93] | <0.001 | 2.18[1.59,2.99] | <0.001 |
| 45-49 | 2.54[1.90,3.40] | <0.001 | 2.52[1.86,3.42] | <0.001 |
| 15-19(RC) | 1 |  | 1 |  |
| **Division** |  |  |  |  |
| Barishal | 1.43[0.96,2.15] | 0.082 | 1.44[0.96,2.15] | 0.078 |
| Chattogram | 1.99[1.29,3.08] | 0.002 | 2.04[1.29,3.22] | 0.002 |
| Dhaka | 1.24[0.84,1.82] | 0.282 | 1.25[0.85,1.85] | 0.258 |
| Khulna | 2.18[1.51,3.15] | <0.001 | 2.17[1.49,3.16] | <0.001 |
| Rajshahi | 2.51[1.71,3.68] | <0.001 | 2.44[1.66,3.58] | <0.001 |
| Rangpur | 1.55[1.02,2.36] | 0.039 | 1.61 [1.06, 2.45] | 0.025 |
| Sylhet | 1.31[0.89,1.93] | 0.167 | 1.36[0.92,2.00] | 0.122 |
| Mymensingh (RC) | 1 |  | 1 |  |
| **Place of residence** |  |  |  |  |
| Rural | 1.09[0.87,1.36] | 0.473 | 0.98[0.78,1.22] | 0.842 |
| Urban | 1 |  | 1 |  |
| **Education of the respondent** |  |  |  |  |
| No education | 1.46[1.13,1.89] | 0.004 | 1.05[1.78,1.43] | 0.737 |
| Primary | 1.29[1.02,1.63] | 0.031 | 1.11[0.87,1.41] | 0.412 |
| Secondary | 1.17[0.95,1.44] | 0.129 | 1.16[0.95,1.41] | 0.153 |
| Higher (RC) | 1 |  | 1 |  |
| **Wealth** |  |  |  |  |
| Poorest | 0.96[0.73,1.25] | 0.74 | 1.04[0.77,1.40] | 0.812 |
| Poorer | 1.18[0.94,1.47] | 0.16 | 1.18[0.93,1.50] | 0.167 |
| Middle | 1.13[0.92,1.39] | 0.243 | 1.12[0.90,1.39] | 0.311 |
| Richer | 0.99[0.81,1.21] | 0.897 | 1.00[0.82,1.22] | 0.979 |
| Richest (RC) | 1 |  | 1 |  |
| **Working status** |  |  |  |  |
| Yes | 1.11[0.97,1.26] | 0.127 | 0.96[0.83,1.12] | 0.630 |
| No (RC) | 1 |  | 1 |  |
| **Has an account in a bank or other financial institution** |  |  |  |  |
| Yes | 1.22[1.05,1.41] | 0.009 | 1.18[1.01,1.38] | 0.042 |
| No (RC) | 1 |  | 1 |  |
| **Participation in decision-making** |  |  |  |  |
| Yes | 1.12[0.98,1.28] | 0.099 | 1.07[0.93,1.22] | 0.953 |
| No (RC) | 1 |  | 1 |  |
| **Attitude toward wife beating** |  |  |  |  |
| Positive | 1.31 [1.12,1.53] | 0.001 | 1.24[1.06,1.45] | 0.008 |
| Negative (RC) | 1 |  | 1 |  |
| **Husband's level of education** |  |  |  |  |
| No education | 1.35 [1.09,1.67] | 0.005 | 1.08[0.87,1.35] | 0.480 |
| Primary | 1.02 [0.84,1.24] | 0.838 | 0.91[0.74,1.12] | 0.373 |
| Secondary | 1.04 [0.87,1.25] | 0.652 | 0.98[0.82,1.16] | 0.804 |
| Higher (RC) | 1 |  | 1 |  |
| **Religion** |  |  |  |  |
| Muslim | 1.31 [0.95,1.81] | 0.105 | 1.35[0.97,1.87] | 0.932 |
| Others (RC) | 1 |  | 1 |  |
| **Exposed to any mass media** |  |  |  |  |
| Yes | 0.97[0.84,1.12] | 0.666 | 0.99[0.87,1.13] | 0.932 |
| No (RC) | 1 |  | 1 |  |
| **Ever used internet** |  |  |  |  |
| No | 1.12[0.96,1.30] | 0.156 | 0.99[0.84,1.19] | 0.986 |
| Yes (RC) | 1 |  | 1 |  |

RC= Reference category

**Table 3** presents factors associated with the treatment of anxiety and depression among participants using log-binomial regression. Women of higher age groups had a higher prevalence ratio of getting treatment than younger age groups. Compared with other age groups, women aged 40-44 (PR:2.21, CI: 1.25, 3.91, p=0.007) and 45-49 (PR:2.11, CI: 1.21, 3.68, p=0.009) had significantly higher prevalence. Women living in Khulna (PR=0.57, CI:0.36, 0.90, p=0.017) had a significantly lower prevalence of treatment of anxiety and depression than other divisions. Women who had an account in a bank or other financial institution (PR=1.38, CI: 1.07, 1.79, p=0.014) had a significantly higher prevalence of treatment than women who didn’t have one. Lastly, women with a positive attitude towards wife beating had significantly higher levels of treatment (PR=1.50, CI: 1.17,1.92, p=0.001) than women with a negative attitude.

**Table 3:** Factors associated with treatment of anxiety and depression using log-binomial regression.

| Variables | Unadjusted model |  | Adjusted model |  |
| --- | --- | --- | --- | --- |
|  | Prevalence ratio [95% CI] | p-value | Prevalence ratio [95% CI] | p-value |
| **Age of the respondent** |  |  |  |  |
| 20-24 | 0.69[0.37,1.30] | 0.256 | 0.72[0.38,1.34] | 0.299 |
| 25-29 | 1.28[0.74,2.20] | 0.372 | 1.32[0.75,2.32] | 0.330 |
| 30-34 | 1.33[0.75,2.37] | 0.325 | 1.35[0.75,2.41] | 0.316 |
| 35-39 | 1.65[0.94,2.88] | 0.082 | 1.69[0.96,2.97] | 0.070 |
| 40-44 | 2.11[1.22,3.64] | 0.007 | 2.21[1.25,3.91] | 0.007 |
| 45-49 | 2.07[1.19,3.60] | 0.010 | 2.11[1.21,3.68] | 0.009 |
| 15-19(RC) | 1 |  | 1 |  |
| **Division** |  |  |  |  |
| Barishal | 1.07[0.69,1.69] | 0.753 | 1.02[0.65,1.60] | 0.942 |
| Chattogram | 1.40[0.90,2.19] | 0.138 | 1.34[0.84,2.13] | 0.222 |
| Dhaka | 0.82[0.51,1.31] | 0.407 | 0.77[0.47,1.28] | 0.320 |
| Khulna | 0.65[0.42,1.02] | 0.062 | 0.57[0.36,0.90] | 0.017 |
| Rajshahi | 1.29[0.85,1.96] | 0.232 | 1.22[0.79,1.88] | 0.359 |
| Rangpur | 0.78[0.49,1.24] | 0.284 | 0.96 [0.57, 1.60] | 0.312 |
| Sylhet | 1.04[0.63,1.73] | 0.873 | 1.36[0.92,2.00] | 0.864 |
| Mymensingh (RC) | 1 |  | 1 |  |
| **Place of residence** |  |  |  |  |
| Rural | 1.22[0.91,1.63] | 0.188 | 1.33[0.93,1.90] | 0.121 |
| Urban | 1 |  | 1 |  |
| **Education of the respondent** |  |  |  |  |
| No education | 1.24[0.81,1.90] | 0.322 | 1.00[0.58,1.73] | 0.999 |
| Primary | 1.26[0.87,1.82] | 0.219 | 1.21[0.75,1.96] | 0.441 |
| Secondary | 1.09[0.76,1.57] | 0.636 | 1.12[0.74,1.70] | 0.603 |
| Higher (RC) | 1 |  | 1 |  |
| **Wealth** |  |  |  |  |
| Poorest | 0.78[0.54,1.13] | 0.188 | 0.80[0.49,1.30] | 0.362 |
| Poorer | 0.89[0.65,1.23] | 0.479 | 0.87[0.58,1.30] | 0.490 |
| Middle | 0.77[0.55,1.07] | 0.123 | 0.75[0.51,1.10] | 0.136 |
| Richer | 0.94[0.68,1.30] | 0.699 | 0.95[0.67,1.34] | 0.757 |
| Richest (RC) | 1 |  | 1 |  |
| **Working status** |  |  |  |  |
| Yes | 0.93[0.73,1.21] | 0.606 | 0.89[0.68,1.16] | 0.385 |
| No (RC) | 1 |  | 1 |  |
| **Has an account in a bank or other financial institution** |  |  |  |  |
| Yes | 1.52[1.21,1.92] | <0.001 | 1.38[1.07,1.79] | 0.014 |
| No (RC) | 1 |  | 1 |  |
| **Participation in decision-making** |  |  |  |  |
| Yes | 0.88[0.70,1.10] | 0.248 | 0.82[0.65,1.02] | 0.076 |
| No (RC) | 1 |  | 1 |  |
| **Attitude toward wife beating** |  |  |  |  |
| Positive | 1.51 [1.18,1.92] | 0.001 | 1.50[1.17,1.92] | 0.001 |
| Negative (RC) | 1 |  | 1 |  |
| **Husband's level of education** |  |  |  |  |
| No education | 1.15 [0.82,1.61] | 0.416 | 1.00[0.64,1.57] | 0.996 |
| Primary | 0.87 [0.62,1.24] | 0.448 | 0.84[0.52,1.34] | 0.460 |
| Secondary | 1.10 [0.80,1.52] | 0.582 | 1.06[0.71,1.58] | 0.772 |
| Higher (RC) | 1 |  | 1 |  |
| **Religion** |  |  |  |  |
| Muslim | 0.86 [0.51,1.46] | 0.584 | 0.87[0.51,1.48] | 0.604 |
| Others (RC) | 1 |  | 1 |  |
| **Exposed to any mass media** |  |  |  |  |
| Yes | 1.08[0.85,1.37] | 0.52 | 1.10[0.85,1.42] | 0.487 |
| No (RC) | 1 |  | 1 |  |
| **Ever used internet** |  |  |  |  |
| No | 0.98[0.76,1.26] | 0.856 | 0.97[0.70,1.34] | 0.852 |
| Yes (RC) | 1 |  | 1 |  |

RC= Reference category

**Table 4** presents factors associated with care-seeking of anxiety and depression among participants using log-binomial regression. Women aged 25 or higher had significant differences with women aged 15-19 in their prevalence of care-seeking. Compared to women aged 15-19, the prevalence of care seeking was significantly higher among those aged 25-29 (PR:1.31, CI: 1.04, 1.66, p=0.027), 30-34 (PR:1.40, CI: 1.10,1.77, p=0.006), 35-39 (PR:1.61, CI: 1.26, 2.06, p<0.001), 40-44 (PR:1.65, CI: 1.27, 2.14, p<0.001), and 45-49 (PR=1.85, CI: 1.37, 2.49, p<0.001). Women living in Barishal (PR=0.60, CI: 0.45, 0.81, p=0.001) had a lower prevalence of care-seeking. However, women living in Chattogram (PR=1.35, CI: 1.07,1.72 p=0.012), Khulna (PR=1.45, CI: 1.17,1.80, p=0.001), and Rajshahi (PR=1.27, CI: 1.00, 1.62, p=0.048) had a significantly higher prevalence of care seeking of anxiety and depression than other divisions. Furthermore, women belonging to middle-class households (PR=1.19, CI: 1.00, 1.41, p=0.046) had a significantly higher prevalence of seeking care than women of other wealth statuses. Women who were working (PR=1.22, CI: 1.09, 1.38, p=0.001) and had a positive attitude toward wife beating (PR=1.52, CI: 1.34, 1.72, p<0.001) received significantly higher care-seeking prevalence. Finally, women who didn’t use the internet had significantly lower care-seeking (PR=0.67, CI: 0.59, 0.77, p<0.001) than those who used the internet.

**Table 4:** Factors associated with care-seeking anxiety and depression using log-binomial regression.

| Variables | Unadjusted model |  | Adjusted model |  |
| --- | --- | --- | --- | --- |
|  | Prevalence ratio [95% CI] | p-value | Prevalence ratio [95% CI] | p-value |
| **Age of the respondent** |  |  |  |  |
| 20-24 | 1.12[0.87,1.43] | 0.372 | 1.14[0.89,1.46] | 0.300 |
| 25-29 | 1.28[1.02,1.61] | 0.034 | 1.31[1.04,1.66] | 0.027 |
| 30-34 | 1.34[1.07,1.68] | 0.01 | 1.40[1.10,1.77] | 0.006 |
| 35-39 | 1.45[1.15,1.84] | 0.002 | 1.61[1.26,2.06] | <0.001 |
| 40-44 | 1.41[1.12,1.78] | 0.003 | 1.65[1.27,2.14] | <0.001 |
| 45-49 | 1.47[1.13,1.90] | 0.004 | 1.85[1.37,2.49] | <0.001 |
| 15-19(RC) | 1 |  | 1 |  |
| **Division** |  |  |  |  |
| Barishal | 0.62[0.46,0.84] | 0.002 | 0.60[0.45,0.81] | 0.001 |
| Chattogram | 1.45[1.15,1.82] | 0.002 | 1.35[1.07,1.72] | 0.012 |
| Dhaka | 1.10[0.86,1.42] | 0.434 | 1.02[0.78,1.33] | 0.893 |
| Khulna | 1.59[1.28,1.97] | <0.001 | 1.45[1.17,1.80] | 0.001 |
| Rajshahi | 1.42[1.11,1.81] | 0.005 | 1.27[1.00,1.62] | 0.048 |
| Rangpur | 1.25[0.98,1.59] | 0.07 | 1.23 [0.97, 1.57] | 0.084 |
| Sylhet | 0.94[0.73,1.21] | 0.639 | 0.96[0.75,1.23] | 0.726 |
| Mymensingh (RC) | 1 |  | 1 |  |
| **Place of residence** |  |  |  |  |
| Rural | 1.09[0.93,1.29] | 0.296 | 1.10[0.93,1.31] | 0.255 |
| Urban | 1 |  | 1 |  |
| **Education of the respondent** |  |  |  |  |
| No education | 0.78[0.63,0.95] | 0.015 | 0.80[0.61,1.04] | 0.089 |
| Primary | 0.92[0.78,1.07] | 0.287 | 1.01[0.83,1.24] | 0.902 |
| Secondary | 1.01[0.87,1.17] | 0.922 | 1.13[0.95,1.34] | 0.178 |
| Higher (RC) | 1 |  | 1 |  |
| **Wealth** |  |  |  |  |
| Poorest | 0.89[0.74,1.07] | 0.197 | 1.24[0.99,1.53] | 0.051 |
| Poorer | 0.93[0.79,1.10] | 0.415 | 1.14[0.95,1.38] | 0.159 |
| Middle | 1.04[0.88,1.22] | 0.672 | 1.19[1.00,1.41] | 0.046 |
| Richer | 0.98[0.84,1.15] | 0.808 | 1.10[0.94,1.28] | 0.259 |
| Richest (RC) | 1 |  | 1 |  |
| **Working status** |  |  |  |  |
| Yes | 1.27[1.14,1.42] | <0.001 | 1.22[1.09,1.38] | 0.001 |
| No (RC) | 1 |  | 1 |  |
| **Has an account in a bank or other financial institution** |  |  |  |  |
| Yes | 1.29[1.15,1.44] | <0.001 | 1.07[0.95,1.21] | 0.263 |
| No (RC) | 1 |  | 1 |  |
| **Participation in decision-making** |  |  |  |  |
| Yes | 1.02[0.91,1.13] | 0.78 | 0.96[0.86,1.07] | 0.429 |
| No (RC) | 1 |  | 1 |  |
| **Attitude toward wife beating** |  |  |  |  |
| Positive | 1.51 [1.33,1.71] | <0.001 | 1.52[1.34,1.72] | <0.001 |
| Negative (RC) | 1 |  | 1 |  |
| **Husband's level of education** |  |  |  |  |
| No education | 0.82 [0.70,0.96] | 0.014 | 0.84[0.70,1.02] | 0.074 |
| Primary | 0.86 [0.74,0.99] | 0.033 | 0.87[0.74,1.03] | 0.110 |
| Secondary | 0.89 [0.78,1.02] | 0.09 | 0.88[0.76,1.03] | 0.114 |
| Higher (RC) | 1 |  | 1 |  |
| **Religion** |  |  |  |  |
| Muslim | 1.02 [0.80,1.32] | 0.859 | 1.05[0.81,1.36] | 0.709 |
| Others (RC) | 1 |  | 1 |  |
| **Exposed to any mass media** |  |  |  |  |
| Yes | 1.15[1.02,1.28] | 0.017 | 1.08[0.96,1.21] | 0.210 |
| No (RC) | 1 |  | 1 |  |
| **Ever used internet** |  |  |  |  |
| No | 0.72[0.65,0.81] | <0.001 | 0.67[0.59,0.77] | <0.001 |
| Yes (RC) | 1 |  | 1 |  |

RC= Reference category

## Discussion

This study explored the situation of diagnosis, treatment, and care-seeking of anxiety and depression and their determinants for reproductive-aged (15-49) women in Bangladesh. Only 2.03% of respondents took medication or treatment provided by a doctor or health care worker. Among all respondents, 3.39% were diagnosed with anxiety, and 0.56% were diagnosed with depression by the provider. Only 7.86% sought help from professionals (doctor/medical personnel), and most of the care was sought from family members, current or former spouses or partners, and neighbors. The diagnosis of mental disorders has been reported to be a stigmatized issue in studies around the world, with a lower rate of diagnosis and reluctance to it.[24–28] In Nepal, only 18.3% of women with any mental disorders (anxiety or depression) sought help[25], and in Ethiopia, 12.9% of women sought help and 4.2% accessed the treatment.[26] Like our findings, women sought most help from informal sources (especially husbands) in Ethiopia.[26] Further, a study of perinatal rural women in Bangladesh found that two-thirds of women didn’t seek any care during that critical period.[17] A study of urban China presented a contradictory picture, with almost 82.2% of women seeking online support for their mental health. [28]

Women’s age, division of residence, and attitude towards wife beating were found to be significant determinants of diagnosis of anxiety and depression. A study about perimenopausal women found higher vulnerability diagnoses of anxiety and depression among this age group.[29] This is consistent with our study as we found higher age as a significant determinant of anxiety and depression. In coherence with our study, domestic violence was found to be a more significant factor in the diagnosis of mental health illness than others.[30–32] A study about postpartum women in Bangladesh reported women suffering from physical, sexual, and psychological abuse suffered from significantly higher amounts of postpartum depression than women who didn’t face any violence.[33]

Furthermore, age, division, accounts in bank or financial institutions, and attitude towards wife beating were significant determinants of treatment. Similarly, a study consisting of a rural Australian sample found that older current age was significantly associated with delayed treatment of anxiety and depressive disorders. [34] A study in Nepal found age and administrative division to be significant factors in mental health treatment among participants.[35]Again, age, division, wealth status, working status, attitude towards wife beating, and internet use were significantly associated with care-seeking. Likewise, in our study, patients’ age was a significant factor in seeking care in a longitudinal study in Sweden. [36]

Our study found that middle-class households were more likely to seek care than other social classes. A study in Nepal also found the same result.[25] The study also found a positive relationship between women’s empowerment and care-seeking for mental disorders, as found in our study. [25] A scoping review also found women’s empowerment incentive as a key to the treatment and care of mental health disorders.[37] In contrast to our study, domestic and intimate partner violence was identified as a barrier to treatment and care-seeking of mental health disorders in a number of studies.[38–40]. Similar to our findings, internet use was found to be an enabling instrument for mental health care seeking and treatment for studies in many countries.[28,41,42]

### Strengths and limitations

It is the first study to explore the situation and factors associated with diagnosis, treatment, and care-seeking among a nationally representative sample of reproductive-aged women. Consequently, it will have better acceptability and generalization. Moreover, our study observed diagnosis, treatment, and care-seeking separately, giving a precise picture of each indicator. This study used log-binomial regression, which is better suited for communication than logistic regression.[23] In addition to strengths, our study has some pivotal limitations. DHS used a cross-sectional research design, which is not exemplary for causal inferences.[43] Additionally, self-reported data may face recall and social desirability bias.[44]

## Conclusion

The diagnosis, treatment, and care-seeking of anxiety and depression among women is still very low in Bangladesh despite women’s unique needs and vulnerable position. Though the overall situation worldwide is not satisfactory, other low-income countries like Nepal and Ethiopia have higher rates of care-seeking and treatment for mental health disorders than Bangladesh. It is also concerning that most care-seeking was done through informal channels. In this situation, the country needs to actively work toward reducing the stigma towards asking for and receiving help for common mental health disorders and create a specialized department in government hospitals for treating anxiety and depression. An awareness program should be designed to reach women of every social class. In addition to women, consciousness needs to build up in society to improve the situation. The internet can be used effectively to increase women’s care-seeking and treatment, as social media use is becoming increasingly common among the overall population of Bangladesh. Special focus should be given to women suffering from intimate partner violence.

## Data Availability

All data produced are available online at https://dhsprogram.com/data/dataset/Bangladesh_Standard-DHS_2022.cfm?flag=0

## Acknowledgements

We would like to thank NIPORT for conducting the Demographic and Health Surveys regularly in Bangladesh.

